# Protocol for a scoping review for risk prediction models for identifying late-onset cardiotoxic effects in cancer survivors for use in a primary care setting

**DOI:** 10.1101/2025.08.13.25333626

**Authors:** Oisín Brady Bates, Alexandru Nicholas Grecu, Jessica Emery, Diarmuid Stokes, Walter Cullen, Joe Gallagher

## Abstract

**Rationale:** After completing treatment for cancer, survivors may experience late cardiotoxic effects: consequences of treatment that persist or arise after a latent period. Primary care providers are well situated to manage these effects. Risk prediction models may assist with this.

**Purpose:** To identify and describe all models that predict the risk of late effects and could be used in an ambulatory care setting.

**Data sources:** Searches will be conducted across MEDLINE and EMBASE supplemented by manual reference list reviews and targeted grey literature searches using Google.

**Study Selection:** Studies to be included are those describing models that 1) predicted the absolute risk of a late cardiotoxic effect present at least one year post-treatment, across all cancer treatment modalities, and 2) could be used in an ambulatory care clinical setting,

**Data Extraction & Synthesis:** Data will be extracted as per the CHARMS checklist, an assessment of bias will be conducted as per the McGinn Checklist.

**Discussion:** This scoping review will comprehensively map existing tools for risk prediction of the late cardiotoxic effects of cancer therapy in cancer survivors, applicable to an ambulatory care setting.

**Registration:** This protocol was registered on 21.6.25 to Zenodo : **10.5281/zenodo.15708760**

**Key Messages:**

1. Cancer survivors face a significant long-term risk of late-onset cardiotoxicity, often undetected until advanced stages.
2. This review will identify and map validated multivariable risk prediction models that could support the early identification of cardiotoxicity in ambulatory and primary care settings.
3. Integrating prediction tools into primary care may help address growing oncology workforce gaps while enabling timely, risk-adapted cardiovascular follow-up.

## Introduction

Cancer therapy-related cardiotoxicity is increasingly recognised as a clinically significant consequence of cancer treatment. A wide range of therapies, including anthracyclines, HER2-targeted agents, radiotherapy, and immune checkpoint inhibitors, have been implicated in both acute and delayed cardiovascular complications (1-3). While some adverse effects emerge during treatment, others, such as heart failure and radiation-induced cardiomyopathy, may not become apparent until several years after therapy has concluded. As cancer survival rates improve, these late-onset cardiac effects are becoming more visible. In the United States, the five-year survival rate for all cancers rose from 60%in 1990 to 71.7% in 2016 (4). With increased longevity, the cumulative burden of cardiovascular disease also rises. This is particularly important given the high prevalence of age-related conditions such as hypertension, diabetes, and coronary artery disease among survivors (5).

The long-term cardiac risks associated with cancer treatment have been well established in paediatric oncology, where decades of survivorship data have documented the association between childhood cancer therapies and subsequent cardiac dysfunction (6-8). Similar trends are now increasingly observed in adults (2, 9). Among breast cancer survivors aged 50 years and older, cardiovascular disease accounts for approximately 35 percent of non-cancer deaths (10). This elevated risk tends to become apparent around seven years following the initial cancer diagnosis (11). Other populations, such as survivors of non-Hodgkin lymphoma and patients treated with tyrosine-kinase inhibitors, have also been shown to experience higher rates of heart failure and left ventricular dysfunction (12, 13).

Late cardiotoxic effects associated with cancer therapies often present sub-clinically and progress silently over years. Risk prediction models can assist in identifying high-risk individuals before overt clinical signs develop. These models are potentially useful at multiple points across the cancer care continuum: prior to treatment initiation, during active therapy, and throughout survivorship (14). At the time of diagnosis, structured cardiovascular risk assessment is recommended. Tools developed by the Heart Failure Association and the International Cardio-Oncology Society provide guidance on stratifying patients based on their risk of treatment-related cardiac complications (2, 15). These assessments typically take into account previous cardiovascular disease, prior cancer treatments, and standard biomarkers such as HbA1c and LDL cholesterol (16). More general tools like the SCORE2 and SCORE2-OP charts may also be used. However, it is important to note that these instruments were not designed specifically for use in oncology populations (15).

Primary care has an important and often under-recognised role to play throughout this continuum. General practitioners are frequently the first point of contact for cancer patients and are well positioned to assess cardiovascular risk, initiate management of modifiable factors, and oversee ongoing surveillance (17, 18). This role becomes especially critical in the post-treatment phase when oncology involvement may decrease but the risk of cardiovascular complications persists (19, 20). However, the transition from specialist-led cancer care to primary care is not always seamless. Many primary care physicians report feeling inadequately trained to monitor for cardiac complications following cancer therapy (21, 22). Moreover, patients with non-specific or ambiguous symptoms may require several visits before appropriate referral takes place (23).

Despite their potential contribution, primary care providers are often underutilised in cardio-oncology. Several barriers have been identified; including the absence of clear guidelines, insufficient training opportunities, fragmented communication between oncology and primary care teams, and patient preferences to remain under specialist care (2, 24-27). Surveys of primary care physicians indicate a strong interest in playing a greater role in cancer care, but many report being engaged too late in the process (28, 29). In addition, conventional cardiovascular risk assessment tools, such as the ASCVD risk calculator, do not account for prior exposure to cardiotoxic cancer therapies. This limits their accuracy in the context of survivorship care (2).

To address this gap, there is a need for validated multivariable risk prediction models that rely on clinical data routinely collected in ambulatory care setting. Variables such as age, blood pressure, cholesterol, and cancer treatment history are readily accessible and could support earlier identification of cardiotoxicity risk (30). Implementation of such tools would enable more timely and effective survivorship care, reduce reliance on specialist services, and strengthen the role of primary and ambulatory care in long-term cardiovascular management for this patient cohort.

### Research Question & Aims

This scoping review will summarise the existing literature relating to risk prediction models with variables that are available in a primary care setting.

The primary research question is:

What are the current validated risk prediction models for identifying late-onset cardiotoxic effects in cancer survivors for use in a primary care setting?

The aims of the review:

- To identify and catalogue existing validated risk prediction models for late-onset cardiotoxicity in cancer survivors
- To describe the study populations and treatment exposures used in model development and validation.
- To determine if current models are suitable for use in ambulatory care settings (hospital outpatient or primary care) (studies of clinical prediction rules conducted in non-primary care settings will be eligible for inclusion if they were relevant to primary care)
- To appraise the cost-effectiveness of utilising these models in identifying cancer survivors for targeted screening for late-onset cardiotoxicity in a cost-effective manner

### Eligibility criteria

Studies will be eligible if they describe risk prediction models for late-onset, cancer-therapy-related, cardiotoxicity, and if they are suitable for use in a primary care setting. The risk prediction model must seek to predict late-onset cardiac toxicity i.e. at least one year after completing therapy, but can seek to predict this at any time from the initial cancer diagnosis to after cancer treatment.

The CHARMS checklist for the evaluation of risk prediction models was used to frame the eligibility criteria. Articles will be excluded if they met at least one of the following criteria: (1) the study was not related to a risk prediction model for late-onset cardiotoxicity in cancer survivors; (2) one or more of the variables in the model are not accessible in anambulatory care setting; (3) the full article could not be obtained; (4) the article was not written in English language; (5) the article wasn’t published in a peer-reviewed journal; and (6) if the article is a review, letter, editorial, or commentary. The inclusion criteria are detailed in Table 1.

**Table 1.**
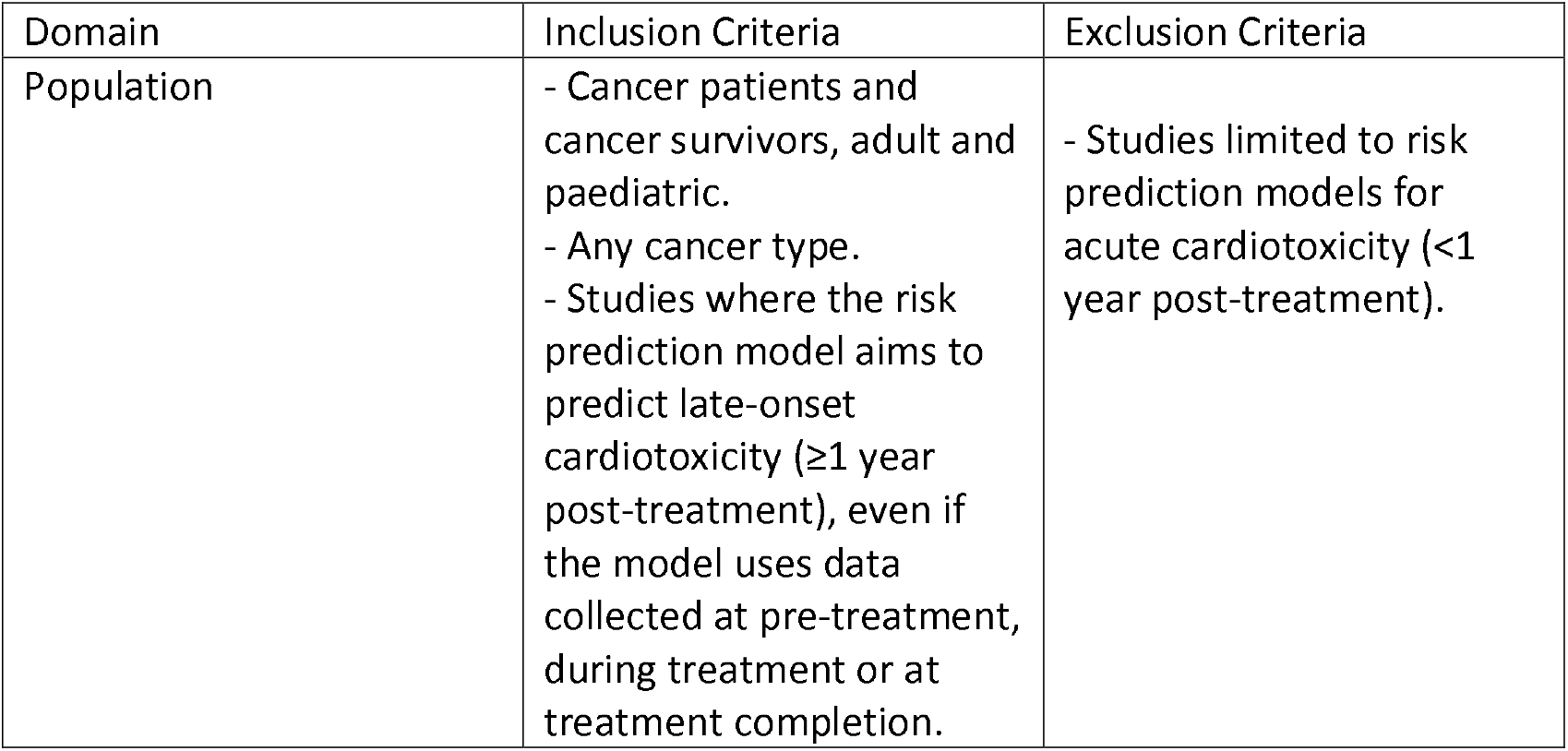
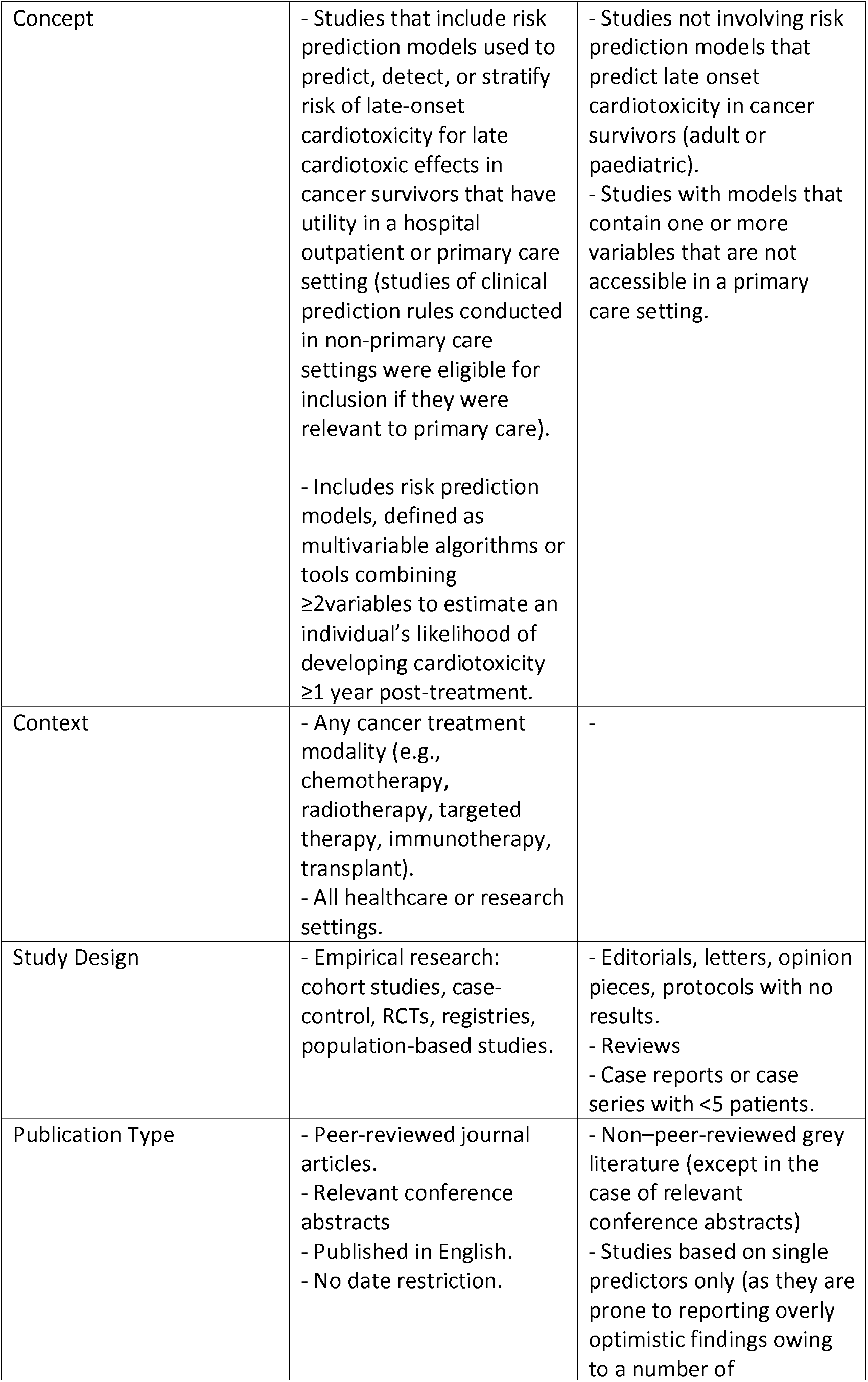
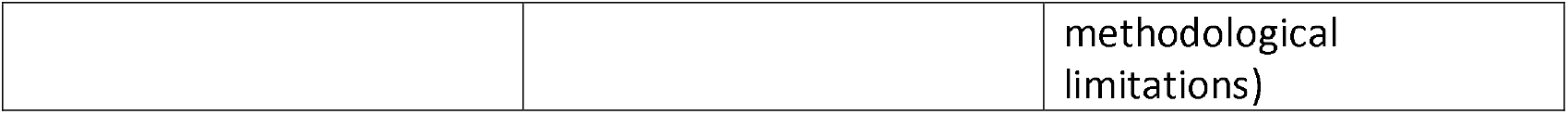
Eligibility Criteria

## Methods

A scoping review was selected as the most appropriate method for this study, as it aims to map the existence of all risk prediction models for the late cardiotoxic effects of cancer therapy, for any cancer type in the literature, providing they contain variables that are available in primary/ambulatory care settings.

The review will adhere to CHARMS checklist (31) for systematic reviews of prediction models and will follow the Preferred Reporting Items for Systematic Reviews and Meta-Analysis extension for scoping reviews (PRISMA-ScR) (32).

The protocol for this review has been registered on Zenodo (**10.5281/zenodo.15708760**).

### Search Strategy

The search strategy will be developed by the entire research team. The review question and design will be framed using the CHARMS checklist for systematic reviews of prediction models (Appendix 1a).

Searches will be conducted across MEDLINE and EMBASE supplemented by manual reference list reviews and targeted grey literature searches using Google. Relevant materials will also be sought from national and international organisations, disease registries, and professional bodies. Only studies published in English will be included, and no date restrictions will be applied. The full search strategy is detailed in Appendix 1b., hosted on the Zenodo.

### Study Selection

The screening process will consist of two stages: (1) title and abstract screening, and (2) full-text review. All citations will be imported into Rayyan (www.rayyan.ai), a Cochrane-endorsed literature screening tool. Two independent reviewers will pilot the inclusion criteria on 10% of abstracts to ensure clarity and consistency before screening the remaining studies. Disagreements will be resolved through discussion or by consulting a third reviewer if necessary. The inclusion criteria are detailed in Table 1. A PRISMA flow diagram will outline the selection process.

### Data Extraction

For each included study, the following information will be extracted on the basis of the CHARMS checklist (Appendix 1c) by O.B.B: source of data, participant characteristics, outcomes to be predicted, candidate predictors, sample size, handling of missing data, model development, model performance (discrimination and calibration), and model evaluation (internal or external validation) and results. Any modifications to the extraction tool during the process will be documented. Authors of included studies may be contacted for missing or additional data. Rayyan will facilitate the extraction process.

The methodological quality of each article will be assessed using criteria based on a previous systematic review of clinical prediction rules in primary care based on modified McGinn criteria (33), in which the criterion concerning ‘100% follow-up’ was changed to ‘adequate follow-up’ and will be defined as ≥80% follow-up of study participants. There are eight criteria assessing internal and external validity for derivation studies, and for validation studies, there are five criteria (Appendix 1d).

### Data Analysis

Findings will be summarized narratively and presented in tables. The CHARMS checklist will be used to critically appraise studies identified and data will be extracted based on the following criteria (31); The CHARMS Framework analysis will guide the categorisation of risk prediction models, biomarker variables utilised within these models, cancer types, treatment modalities and model evaluation methods.

Rigour will be maintained through an audit trail of decisions throughout the review process and through the triangulation of review findings with existing literature and ongoing stakeholder input.

### Study Status

At the time of publication, the review is at the full-text screening stage.

## Discussion

This review will provide a comprehensive overview of existing risk prediction models for late-onset cardiotoxicity in cancer survivors, with a focus on their applicability in primary/ambulatory care. It aims to explore how routinely available data in ambulatory care settings can be integrated into predictive models to support earlier identification of cardiovascular risk and guide survivorship care. The review will also highlight opportunities to strengthen collaboration between primary care and oncology through enhanced education and predictive approaches.

Strengths of this review include its multidisciplinary perspective and attention to real-world applicability. The findings will be disseminated through peer-reviewed publications and presentations at conferences.

## Supporting information

Appendix Items

## Acknowledgements

N/A

## Declarations

### Ethics

Given that the scoping review methodology consists of the examination and collation of information from publicly accessible resources, ethical approval is deemed unnecessary for this study.

### Patient and public involvement

Patients or the public were not involved in the design, or conduct, or reporting, or dissemination plans of our research.

### Transparency statement

The lead author confirms that this manuscript is an honest, accurate, and transparent account of the study being reported; no important aspects of the study have been omitted; and that any discrepancies from the study as originally planned have been explained.

## Availability of data and materials

### Underlying data

No underlying data are associated with this article.

### Extended data

Zenodo: **10.5281/zenodo.15708760**

Data are available under the terms of the Creative Commons Attribution 4.0 International Public License.

## Data Availability statement

This is an article outlining a scoping review protocol. No data is available for sharing.

## Reporting Guidelines

PRISMA SCR Checklist available at Zenodo **10.5281/zenodo.15708760**

