## Appendix Items for "Protocol for a scoping review for risk prediction models for identifying late-onset cardiotoxic effects in cancer survivors for use in a primary care setting"

### **Appendix 1a CHARMS key items**

Design of the systematic review on diagnostic prediction models for the diagnosis of late-onset cardiotoxicity in adult cancer patients based on the CHARMS.

| Item |  |
| --- | --- |
| **1. Scope of the review** | Review of all existing prediction tools that utilise multivariable models for the prediction of late onset cardiotoxicity in adult cancer survivors.  Risk prediction models can be defined as multivariable algorithms or tools combining at least two variables to estimate an individual’s likelihood of developing cardiotoxicity >1 year post-treatment.  Studies based on single predictors only will be excluded (as they are prone to reporting overly optimistic findings owing to a number of methodological limitations).  The review will exclude models limited to the diagnosis of acute cardiotoxicity i.e. patients admitted to hospital or attending a hospital/primary care healthcare setting with acute toxicity >1 year post treatment. |
| **2. Type of primary studies** | Diagnostic prediction model development studies with or without external validation in independent data. |
| **3. Target population** | Risk prediction models must be for use with adult cancer patients or survivors (≥18 years) who have completed cancer treatment. Any cancer type will be included. |
| **4. Outcome to be predicted** | Incident cardiotoxicity present or absent as determined by an established reference standard, such as echocardiography, cardiac MRI, specialist opinion using reference criteria e.g. Framingham or a combination of these. |
| **5. Intended utility of the model** | For the diagnosis of late cardiotoxic effects of cancer treatment in adult cancer survivors. |
| **6. Model setting of care** | Hospital outpatient or primary care (studies of clinical prediction rules conducted in non‐primary care settings are eligible for inclusion if they are relevant to primary care). |

### **Appendix 1b Search strategy**

Pubmed

#1 "primary care" OR "Primary Health Care" OR “primary medical care” OR "general practice" OR "family practice" OR "general practise" OR "family practise" OR "ambulatory care" OR "community setting" OR "outpatient*" OR "Primary Health Care"[Mesh] OR "Outpatients"[Mesh] OR "Ambulatory Care"[Mesh:NoExp] OR "General Practice"[Mesh]

AND

#2 "risk prediction model*" OR "Clinical Decision Rules" OR "Risk Factors" OR "clinical prediction rule*" OR "risk score*" OR "prognostic model*" OR "multivariable model*" OR "decision support tool*" OR "Clinical Decision Rules"[Mesh] OR "Risk Factors"[Mesh]

AND

#3 cardiotoxic* OR "cardiac dysfunction" OR "cardiac toxicity" OR "cardiovascular complication*" OR "heart disease*" OR "heart failure*" OR "Cardiovascular Diseases"[Mesh]

#4= #2 AND #3

AND

#5 Cancer* OR Neoplasm* OR oncology OR Neoplasia* OR Tumor* OR Tumour* OR Malignanc* OR "Neoplasms"[Mesh]

AND

#6 survive* OR survival OR survivor* OR "Living with" OR "post treatment" OR "after treatment" OR recover* OR "Survivors"[Mesh:NoExp]

#7 =#5 AND #6

#8 "Cancer Survivors"[Mesh]

#9= #7 OR #8

#10 = #1 AND #4 AND #9

Embase

#1 "primary care" OR "Primary Health Care" OR 'primary medical care' OR "general practice" OR "family practice" OR "general practise" OR "family practise" OR "ambulatory care" OR "community setting" OR "outpatient*" OR 'primary medical care'/de OR 'general practice'/de OR 'ambulatory care'/de OR 'outpatient'/de

AND

#2 "risk prediction model*" OR "clinical prediction rule*" OR 'clinical decision rule' OR "risk score*" OR "prognostic model*" OR "multivariable model*" OR "decision support tool*" OR 'clinical decision rule'/de

AND

#3 cardiotoxic* OR "cardiac dysfunction" OR "cardiac toxicity" OR "cardiovascular complication*" OR "heart disease*" OR "heart failure*" OR " 'cardiotoxicity'/de OR 'cardiotoxin'/de OR 'cardiovascular disease'/exp OR 'heart failure'/exp

#4= #2 AND #3

AND

#5 Cancer* OR Neoplasm* OR oncology OR Neoplasia* OR Tumor* OR Tumour* OR Malignanc* OR 'malignant neoplasm'/exp

AND

#6 survive* OR survival OR survivor* OR "Living with" OR "post treatment" OR "after treatment" OR recover* OR 'survivor'/de OR 'survival'/de OR 'cancer survival'/exp OR 'post treatment survival'/de

#7 =#5 AND #6

#8 OR 'cancer survivor'/de

#9= #7 OR #8

#10 = #1 AND #4 AND #9

**Appendix 1c CHARMS Checklist Extraction Tool**

Source of data

Participants

Outcomes to be predicted

Candidate predictors

Sample size

Missing data

Model development

Model performance

Model evaluation

Results

Final variables

### **Appendix 1d Assessment of bias**

| Derivation | All-important predictors included? | All-important predictors in a significant proportion | Outcome events and predictors clearly defined | Outcome event assessors blinded | Predictor event assessors blinded | Sample size adequate? | Clinically sensible? |
| --- | --- | --- | --- | --- | --- | --- | --- |
| Validation | **Participants chosen in unbiased fashion** | **Represent a wide variety of severity of disease** | **Blinded assessment of criterion standard** | **Adequate follow-up** | **Explicit and accurate interpretation of rule** |  |  |
